# Idiopathic and acquired pedophilia as two distinct disorders: an insight from neuroimaging

**DOI:** 10.1101/2020.06.26.20140798

**Authors:** Cristina Scarpazza, Livio Finos, Sarah Genon, Laura Masiero, Elena Bortolato, Camilla Cavaliere, Jessica Pezzaioli, Merylin Monaro, Nicolò Navarin, Battaglia Umberto, Pietro Pietrini, Stefano Ferracuti, Giuseppe Sartori, Andrea S. Camperio Ciani

## Abstract

**Objective:** Pedophilia is a disorder of public concern because of its association with child sexual offense and recidivism. Previous neuroimaging studies identified inconsistent brain abnormalities underlying pedophilic behavior both in idiopathic and acquired (i.e., emerging following brain damage) pedophilia. This study sought to explore the neural underpinnings of pedophilic behavior and to determine the extent to which brain alterations may be related to distinct psychopathological features in pedophilia.

**Methods:** A coordinate based meta-analysis on previously published papers reporting whole brain analysis and a lesion network analysis using brain lesions as seeds in a resting state connectivity analysis were run to investigate the presence of consistent neural basis of idiopathic and acquired pedophilic behavior, respectively. The behavioral profiling approach was applied to link identified regions with corresponding psychological processes.

**Results:** While no consistent neuroanatomical alterations have been identified in idiopathic pedophilia, the current results support that all the lesions causing acquired pedophilia localized to a shared resting state network including posterior midlines structures, right inferior temporal gyrus and bilateral orbitofrontal cortex. These regions are associated with action inhibition and social cognition, abilities that are consistently described as severely impaired in acquired pedophiles.

**Conclusions:** This study suggests that idiopathic and acquired pedophilia may be two distinct disorders, in line with their distinctive clinical features, including age of onset, reversibility and modus operandi. Understanding the neurobiological underpinnings of pedophilic behavior may contribute to a more comprehensive characterization of these individuals on a clinical ground and to develop more efficient therapeutic rehabilitation strategies.

## Introduction

Pedophilia is a paraphilic disorder included within the Diagnostic and Statistic manual of Mental Disorder (fifth edition^1^) defined as sexual preferences for prepubescent children, coupled with distress caused by the sexual urges and/or child sexual abuse^1, 2^. Although pedophiles are relatively rare (prevalence of 3% to 5% in the male population^3^), they commit a disproportionate amount of crimes and rarely comply with psychological treatments^4^. Pedophilia raises high public concern due to its association with child sexual offense and recidivism^5^.

Though the multifactorial origin of pedophilia still remains elusive ^6, 7^, recent neuroimaging studies have shown pedophilia to be associated with reduced grey^8^ and white^9^ matter in brain regions involved in sexual arousal^6^, including amygdale, hypothalamus and septal regions, as well as in orbitofrontal cortex (OFC) and basal ganglia. Pedophiles also showed significant functional activation differences while viewing images depicting nude children and nude adults as compared to controls^10^. Overall, studies revealed a widespread dysfunctional brain activity mainly encompassing the frontal, parietal and temporal lobes, as well as relevant subcortical structures.^7^ However, structural and functional abnormalities in pedophiles show considerable variability across studies. Furthermore, it is unclear to what extent these abnormalities are an incidental correlate rather than a cause of pedophilia^6, 7^.

As psychiatric symptoms can emerge as a consequence of neurological insult^11, 12^, an effective approach, typical of classical neuropsychology, to investigate the neural basis of pedophilia is to study patients who develop pedophilic urges and/or behavior following focal brain lesions, referred to as “acquired pedophilia”^13, 14^. Unlike “idiopathic pedophilia”, whose etiology is unknown, acquired pedophilia occurs *de novo* in individuals who had never manifested pedophilic interests or urges earlier in life, as a symptom of an underlying neurological disorder.^15^ The causal inference is strengthened by a detectable temporal relationship between the onset of the neurological disorder and the pedophilic behavior^16^. Furthermore, pedophilic behavior recedes after treating the underlying neurological condition^17, 18^. The first documented case of acquired pedophilia, reported in 1862^19^, was a senescent man without any previous criminal record charged with child abuse as he sexually assaulted an underage boy playing in the garden. Upon medical examination, he manifested memory deficits, tangential language, inability to appreciate the moral disvalue or the legal implications of his behaviors. Eventually, he was diagnosed with dementia. More recent cases of acquired pedophilia reported in the literature include: a middle age man who manifested pedophilia because of a tumor in the OFC^17^; a senescent man who displayed heightened sexual interest toward children after hippocampal sclerosis^20^; a middle age man who began to make sexual proposal to children due to a glioma involving the thalamus, hypothalamus, ventral midbrain an pons^21^. The above examples suffice to understand that mere lesion localization is unsuited to explain the neurological origin of acquired pedophilia, as different cases may implicate different brain regions.

Overall, brain imaging studies in idiopathic and acquired pedophilia are inconclusive, as they show subtle and inconsistent brain alterations in developmental pedophilia, and spatially heterogeneous brain lesions in acquired pedophilia. Furthermore, it is not clear whether or not they share the same anatomical substrate.

The current study aims to: i) identify the brain regions consistently impaired in idiopathic and acquired pedophilia; ii) determine whether the two forms of pedophilia are associated with overlapping or separate brain networks; iii) link topographically defined regions with corresponding psychological processes testing which kind of experiments are most likely to activate a given region, to give a cognitive/psychological meaning to the detected alterations.

In order to identify brain regions consistently impaired in idiopathic pedophilia, a coordinate based meta-analysis using the Activation Likelihood Estimation (ALE) method was performed^22^. This approach reveals converging and consistent findings across different studies, underlying important nodes of network alteration in idiopathic pedophilia.

This strategy cannot be used for acquired pedophilia, as only cases reports, with macroscopic neuroanatomical alterations have been published. Thus, the lesion network mapping approach was used to identify brain regions consistently impaired in acquired pedophilia.^23^ This method, assuming that every brain region is a part of complex network, identifies regions functionally connected to a lesion localization. Indeed, lesions causing the same symptoms tend to be functionally connected with the same brain regions.^23^

## Material and Methods

### Idiopathic Pedophilia

#### Studies Selection and data extraction

An in-depth search was conducted on Pubmed up to January 2020. The following terms were used: (“pedophilia” OR “pedophilic behavior” OR “child* abuse”) AND (“fMRI” OR “functional magnetic resonance imaging” OR “neural basis” OR “voxel based morphometry” OR “brain abnormal*”). A search for studies in review and meta-analysis articles and a reference tracing were also performed.

Studies meeting the following criteria were included: i) using structural (sMRI) or functional (fMRI) MRI; ii) whole brain analysis (i.e., studies performing only region of interest (ROI) analysis were excluded); iii) original peer-reviewed data; iv) including a healthy control group (HC) or investigating neural differences between pedophilic individuals and HC or between pedophilic individuals who committed sexual abuse and pedophilic individuals who did not commit sexual abuse; v) sample size >=5 (per group); vi) results in a standardized coordinate space (e.g., Tailarach or Montreal Neurologic Institute-MNI-).

Literature screening and selection was performed according with the PRISMA guidelines^24^. Two authors (CS and MM) screened the data independently. A third opinion (UB) was sought in case of discordance. Characteristics such as sample size, age of participants, coordinate space, coordinates and statistical values were recorded.

#### Statistical analysis

For a quantitative assessment of inter-study concordance, the Activation Likelihood Estimation (ALE) method^22^ applied to both structural and functional data, following the most recent guidelines^25^.

The peaks of activation/deactivation or of increased/decreased grey matter volume were used to generate an ALE map, using the revised ALE algorithm^26^ running under Ginger ALE software (http://brainmap.org/ale/) version 3.0.2. This algorithm treats activated foci of brain regions as three-dimensional Gaussian probability distributions centered at the given coordinates.^22^ The algorithm incorporates the size of the probability distributions by considering the sample size of each study. Moreover, it employs the random-effect rather than the fixed-effect inference, by testing the above chance clustering between contrasts rather than the above-chance clustering between foci. If the study reported more than one contrast of interest (for instance: brain activation while seeing adult vs child naked body), only the contrast more representative of the process of interest was selected. This procedure was applied to adjust for multiple contrasts from the same sample^25^. Then, inference was sought regarding regions where the likelihood of activation being reported in a particular set of experiments was higher than expected by chance. Tailarach coordinates were transformed in MNI using a linear transformation. Statistical parametric maps were thresholded using cluster level family-wise error (FWE) correction at p<0.05 (cluster-forming threshold at voxel-level p<0.001). For explorative analysis only, a p<0.001 uncorrected threshold was used. The correspondent brain regions were identified using the SPM Anatomy toolbox (version 1.5)^27^.

### Acquired Pedophilia

#### Studies Selection and data extraction

Published cases of acquired pedophilia were identified through a systematic review^13^. Cases meeting the following criteria were included: i) studies describing new cases of late onset pedophilic behavior; ii) pedophilic behavior emerged following a documented neurological condition; iii) pedophilic behavior had a clearly identifiable neural basis. Two authors (CS and UB) extracted and screened the data independently. A third opinion (MM) was sought in case of discordance. Information as age of the offender, etiology of the underlying neurological disorder, brain localization of neuroanatomical abnormalities and symptoms other than pedophilia were recorded.

#### Statistical analysis

The lesion network mapping analysis^23^ was run to determine whether these lesion locations were part of a common brain network.

First, brain alterations causing the onset of pedophilic behavior were identified in each patient and manually traced in consensus by two expert raters (CS and UB) on the axial image of a standardized template using the MRIcron software (available at http://www.mricro.com/mricron).^28^ Then, the lesion outline was verified by an independent third rater (SF). Some patients presenting with *de novo* pedophilia were diagnosed with a behavioral variant of frontotemporal dementia (bvFTD) and, thus, they did not present a spatially defined lesion that could be outlined. In order to identify the neural structures consistently impaired in bvFTD, a coordinate based meta-analysis was run on papers presenting structural or functional abnormalities in patients with bvFTD vs. healthy controls (see Supplementary Material A and B for details). The output of the meta-analysis was then transformed in a binary mask.

Second, traced lesions were used as individual seeds in a seed based connectivity analysis, using resting state fMRI data from one hundred healthy subjects randomly selected from a freely available dataset: https://openneuro.org/datasets/ds000221. The brain functional connectivity with each lesion was determined by calculating the correlated time course between each lesion location and every other brain voxel using the resting-state data from each individual healthy control, as previously reported.^23^ The results in all controls were combined into an average correlation (r), converted according to Fisher transformation (z) using the following formula:

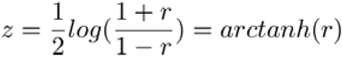

and then modeled in a linear regression framework to obtain a T value for each individual voxel and each brain mask. Voxels were thresholded at T > ±17 to create a binarized map of significantly functionally connected regions to the seed, that corresponded to an effect size of R^2^=0.75. An extent threshold of 50 voxels also was applied. In this way, the brain network impaired by the presence of each lesion was calculated. Finally, maps from each of the patients were combined to form the lesion network mapping overlap for the group, showing the number (and percentage) of patients with lesions functionally connected to each individual voxel. A stability analysis was performed by replicating the analyses using three different control groups, each with 25 healthy subjects (we kept the minimum effect size of R^2^=0.75, which implies a T>±8.5). Analyses were performed using SPM-CONN (2018b) adopting standard preprocessing and denoising steps.

### Behavioral profiling

To link topographically defined brain regions with corresponding psychological processes, we ran a behavioral profiling approach across databases of aggregation of activations experiments.^29^ This approach allows to identify which kind of experiments are most likely to activate a given region. A reverse inference approach with statistical testing (P < .05 corrected for multiple comparisons) was performed on the identified clusters of voxels in BrainMap database (http://www.brainmap.org/) to identify the Behavioral Domain and Paradigm Classes consistently associated with these regions.

## Results

### Idiopathic Pedophilia

One hundred and eighty studies were identified. After excluding the articles that did not meet the inclusion criteria, 19 original articles were included. The screening procedure, summarized in the PRISMA diagram, and reasons for excluding individual studies are reported in the Supplementary Material C.

The included studies are summarized in Supplementary Material C and the full database reporting the coordinates is available in the Supplementary Material D. Briefly, the coordinate based meta-analysis comprises 20 experiments (one study included two independent groups of pedophiles), 240 foci, 436 pedophiles and 449 the control groups, of which 302 are healthy controls, 50 are non sexual offenders, and 97 are pedophiles who did not commit sexual offenses toward children. The included studies are not completely independent as some come from the same laboratories and at least a partial sample overlap is reported in some studies.

Despite the power of the analysis was sufficient to achieve significant results^30^, using a conservative statistical threshold, no significant results were found. For exploratory purposes only, the threshold was decreased to p<0.001 uncorrected. Using this liberal threshold, four clusters located in the middle occipital gyrus (coordinates: -36, -78, 2), in the middle cingulate gyrus (coordinates: 8, -12, 42 and 12, -30, 46) and in the superior frontal gyrus (coordinates: -17, 24, 45) were found (Figure 1). As the foci contributing to each cluster come from independent samples and given the exploratory nature of this second analysis, the meta-analysis was not repeated removing overlapping samples. The behavioral profiling analysis was not performed in order to avoid over-interpretation of statistically non-significant results.

**Figure. 1:** Results of ALE-meta-analysis in idiopathic pedophilia. Results are presented in the sagittal view for illustrative purposes only at the liberal statistical threshold of p<0.001, uncorrected. IPL: Inferior Parietal Lobe; MCC: Middle Cingulate Cortex; IOG: Inferior Occipital Gyrus; SFG = Superior Frontal Gyrus.

### Acquired Pedophilia

Seventeen papers were identified through literature search, for a total of nineteen cases that met our inclusion criteria (see Supplementary Material E for details on included and excluded cases). Seven out of 19 patients expressed hyper-sexuality and all of them manifested a more general impulse dis-control. Moral judgment/social cognition behavior (namely, the ability to understand the social and moral disvalue of their action, theory of mind, ability to discriminate right from wrong) were impaired in nine patients, spared in four, while no data were available for the remaining six cases.

Lesion localization was very heterogeneous, as reported in Supplementary Material E. Lesions were traced using the original anatomical scans of the patients in two cases; the images reported in the original publications in six cases; from a coordinate based meta-analysis on bvFTD in four cases. In the remaining cases, lesions were traced following the description provided in the paper and following indications from expert neuroradiologists and neurosurgeons when needed. In one case, the author of the original publication verified the traced lesion.

The lesion network mapping analysis revealed that, though individual lesions had different locations, 95% of them were part of a single brain network defined by functional connectivity with posterior midline structures (center of gravity coordinates: 0, -43, 55), including the posterior cingulate cortex (PCC) and precuneus; the OFC bilaterally (left coordinates: -34, 32, -17; right coordinates: 36, 30, -17)); the right inferior temporal gyrus (ITG) (coordinates: 52, -16, -28), the left calcarine gyrus (coordinates: -9, -56, 7) and the left fusiform gyrys (coordinates: -44, -63, -19) (Figure 2). These results were replicated also using smaller control groups.

**Figure 2:** Brain regions consistently involved in acquired pedophilia. OFC= OrbitoFrontal Cortex, PCC= Posterior Cingulate Cortex; ITG = Inferior Temporal Gyrus; R = right; L = left

Interestingly, the behavioral profiling analysis (Table 1) highlighted that the identified regions are associated with social cognition (posterior midline structures and inferior temporal gyrus) and in particular with the theory of mind construct (posterior midline structures). Furthermore, a significant association was found between the right OFC and different functions covering emotions and action inhibition. Additionally, regions in the left hemisphere were associated with functions supporting object identification/interpretation, monitoring of information/discrimination judgments and autobiographical remembering.

**Table 1.** Results of the behavioral profiling analysis. PCC = Posterior Cingulate Cortex; OFC = OrbitoFrontal Cortex; IFG = Inferior Temporal Gysus; ns = non significant results.

| **Cluster** | **Size (k)** | **BrainMap Behavioral Domains** | **BrainMap Paradigm Classes** |
| --- | --- | --- | --- |
| Posterior midline regions (precuneus, PCC) | 131 | Social Cognition | Theory of mind |
| Left OFC | 122 | Language Cognition: Semantics, Gustation | Semantic Monitor/discrimination |
| Right OFC | 62 | Emotion, Gustation, Action Inhibition | ns |
| Right ITG | 96 | Social Cognition | ns |
| Left Fusiform gyrus | 95 | Language Cognition: Semantics, Speech, Phonology.<br>Action execution: Speech, Action observation | Face; monitor/discrimination; Phonological discrimination; film viewing; naming (overt); naming (covert) |
| Left Calcarine Gyrus | 55 | Explicit (long-term) memory | Autobiographical recall |

## Discussion

This study sought to: i) identify consistent alterations associated with acquired and idiopathic pedophilia; ii) understand whether and to what extent the two forms of pedophilia may share the same biological substrate; iii) investigate whether consistent brain abnormalities may explain psychopathological features typically detected in pedophiles.

Of relevance, the lesion network mapping technique revealed that the neural bases of acquired pedophilia localize to a common resting state network, despite their high spatial heterogeneity. Overall, these data support a shared neurobiological substrate in acquired pedophilia, as they reveal that the lesions temporally associated with acquired pedophilic behavior are all functionally connected with a network involving the OFC areas, the posterior midline structures, the right inferior temporal gyrus and the left fusiform gyrus. On the contrary, the ALE meta-analysis of whole brain neuroimaging studies in idiopathic pedophilia revealed no spatially convergent findings across studies, suggesting that idiopathic pedophilia is characterized by the absence of consistent alterations that may be detected by structural or functional neuroimaging investigations. However, when lowering the statistical threshold, a few clusters of spatial convergence emerged in the superior frontal gyrus, middle cingulate and middle occipital gyrus, suggesting that idiopathic and acquired pedophilia may have distinct neural bases. Of note, the amygdala, which had been reported to be consistently impaired in pedophilia^6, 7^ did not emerge from the current meta-analysis, even at the more liberal statistical threshold, suggesting that ROI analyses in original studies may have overestimated the amygdala effects.

Despite idiopathic and acquired pedophilia are usually considered as a whole in studies that investigate the neural basis of pedophilic behavior, they actually seem to be two distinct disorders. Indeed, they differ in their etiology: while idiopathic pedophilia is a paraphilia, namely a psychiatric disorder included within the DSM5, acquired pedophilia is a symptom resulting from an underlying neurological insult. Second, their *modus operandi* widely differs: while idiopathic pedophiles are characterized by a highly predatory style^4, 31^, acquired pedophiles usually show a dis-organized behavior, characterized by dis-control of impulses^13, 16^. Third, the temporal insurgence of the pedophilic urges is an additional factor that contributes to the differential diagnosis: while idiopathic pedophilia typically first appears in adolescence and is stable across the lifespan^3^, the age of the onset of acquired pedophilia is variable and typically well after adolescence, due to its acquired origin^13^. The current results further expand previous literature identifying two dissociable neural networks involved in the two forms of pedophilia, corroborating the emerging idea that they might be two different disorders/distinct entities^13^.

Interestingly, the data driven behavioral profiling on acquired pedophilia indicated impaired connectivity between lesions causing acquired pedophilia and regions crucial for social cognition (posterior midline structures and right ITG), in particular the theory of mind (posterior midline structures), emotion recognition (right OFC), impulse control (right OFC), semantic interpretation of cues (left OFC, L fusiform gyrus). It is noteworthy that these results match well with the aberrant behavior pattern described in acquired pedophiles. The observation of altered activity in a key region for impulse inhibition fits perfectly with previous evidence from single case description of patients with acquired pedophilia, in whom dis-inhibition was invariably present^14, 18, 20, 21, 32, 33^. Dis-inhibition also was recently reported to be a red flag suggesting an acquired origin of pedophilic behavior^13^ and explains the dis-organized *modus operandi* of these sexual offenders. Similarly, the observation of altered activity in key regions for social cognition, in particular for theory of mind and emotion recognition, fits well with these patients inability to understand what is morally wrong ^13, 18, 20, 21, 33-36^.

Of note, these results are specific for acquired pedophilia, as idiopathic pedophilia was not associated with impairments in the same brain regions, even when the statistical threshold was lowered. Idiopathic pedophiles are characterized by a different profile of neuropsychological impairment, consisting in a lower IQ and working memory performance, coupled with higher performance in abstract reasoning and planning, as compared to non pedophiles ^6^. Despite difficulties in behavioral inhibition and empathy have also been observed in idiopathic pedophiles, the reported effect size is very small^6^, suggesting that individual inference in idiopathic pedophilia are relevant. Furthermore, idiopathic pedophilia has a high comorbidity with other psychiatric disorder, in particular with personality disorder.^37^ Thus, it is difficult to disentangle whether the reported neuropsychological impairments are related to pedophilia itself or to the associated psychiatric disorder.

Importantly, cognitive abilities associated with brain regions impaired in acquired pedophiles, and found defective in individual patients (impulse control, emotion recognition and social cognition/theory of mind) are pivotal for self-determination. Indeed, the concomitant impairment of impulse control and social cognition, following the neuroscientific INUS (Insufficient but Non-redundant parts of Unnecessary but Sufficient conditions) model of causation^38^, could account for the acquired pedophilic behavior. According to this model, none of these impaired functions by itself alone could explain the insurgence of pedophilic behavior, but all together they are.

Interestingly, according with previous hypotheses, acquired pedophilia may be considered as a behavioral manifestation of pre-existent latent pedophilic urges due to general impulse dis-inhibition^7^. The application of the INUS model of causation to acquired pedophilia, requiring the concomitant presence of both social cognition and action inhibition impairment, suggests that this hypothesis may be ruled out. This claim, however, needs further exploration.

As a final note, the result that idiopathic and acquired pedophilia are characterized by distinct neural networks highlights the need to reconsider the approach of using neurological disorders to investigate the basis of psychiatric conditions or complex behaviors^23^. Indeed, individual psychiatric symptoms that may appear within the clinical picture of a neurological condition, like hallucinations or thought disorders in patients with epilepsy or brain tumors, not necessarily may have a neural substrate identical to the one underlying their manifestation in the course of a psychiatric disorder. As a matter of fact, psychiatric and neurological disorders have been proven to have distinct neuroimaging correlates that arguably may reflect distinct neuropathologies^39^.

This study is not free from drawbacks. In particular, some of the seeds of the lesion network analysis were traced without a 2D figure present in the original paper and required consulting with neuroradiologists to identify the most likely lesion. Results of neuroimaging analysis, however, strongly reflect cognitive /behavioral deficits observed in those patients, corroborating the plausibility of our analysis. Second, the lesion network analysis was run using 100 healthy controls only. The additional analyses we run, however, corroborated the robustness of the results, which remained stable using different control groups of 25 healthy controls.

In summary, the results of this study pinpoint aberrant brain activity related to acquired but not to idiopathic pedophilia. All the lesions causing acquired pedophilia localized to a shared resting state network including the posterior midlines structures, the right inferior temporal gyrus and the bilateral OFC, regions consistently involved in social cognition, theory of mind, emotion recognition and action inhibition. Alterations of these neuropsychological functions have been consistently described in individual reports of acquired pedophiles, supporting the reliability of the data. Interpreting these results in light of the INUS model of causation is relevant to better characterize these patients and plan therapeutic rehabilitation^40^. Further researches are needed to corroborate these results.

## Data Availability

All data used can be provided

## Acknowledgements

CS was supported by a grant from the University of Padua (Supporting TAlent in ReSearch @ University of Padua - STARS Grants 2017). The present study was carried out within the scope of the research program “Dipartimenti di Eccellenza” (art.1, commi 314–337 legge 232/2016), which was supported by a grant from MIUR to the Department of General Psychology, University of Padua.

## Disclosures

The authors reported no biomedical financial interests or potential conflicts of interest.

## Tables Legend

**Table 1.** Characteristics of the studies included in the ALE meta-analysis on idiopathic pedophilia. Ped. N= Number of pedophiles; Contr. N = number of controls; HC = Healthy Controls; NSO = Non Sexual Offenders; CSA+ = pedophiles who committed child sexual abuse; CSA- = pedophiles who did not commit child sexual abuse; sMRI= structural magnetic resonance images; fMRI= functional magnetic resonance images.

**Table 2.** Clinical characteristics of the patients with acquired pedophilia. OFC = OrbitoFrontal Cortex; vmPFC = VentroMedial Prefrontal Cortex; PFC = Prefrontal Cortex; bvFTD = behavioral variant Fronto Temporal Dementia; TBI = Traumatic Brain Injury.

**Table 3.** Results of the behavioral profiling analysis. PCC = Posterior Cingulate Cortex; OFC = OrbitoFrontal Cortex; IFG = Inferior Temporal Gysus; ns = non significant results.

## Notes

### Competing Interest Statement

The authors have declared no competing interest.

### Funding Statement

No external funding was received

## References

1. Association. AP. Diagnostic and statistical manual of mental disorders. Arlington: American Psychiatric Publishing 2013.

2. Regier DA, Kuhl EA and Kupfer DJ. The DSM-5: Classification and criteria changes. World Psychiatry 2013; 12: 92-98. 2013/06/06. DOI: 10.1002/wps.20050.

3. Beech AR, Miner MH and Thornton D. Paraphilias in the DSM-5. Annu Rev Clin Psychol 2016; 12: 383-406. 2016/01/17. DOI: 10.1146/annurev-clinpsy-021815-093330.

4. Hall RC and Hall RC. A profile of pedophilia: definition, characteristics of offenders, recidivism, treatment outcomes, and forensic issues. Mayo Clin Proc 2007; 82: 457-471.2007/04/10. DOI: 10.4065/82.4.457.

5. Hanson RK, Morton KE and Harris AJ. Sexual offender recidivism risk: what we know and what we need to know. Ann N Y Acad Sci 2003; 989: 154-166; discussion 236-146. 2003/07/04.

6. Tenbergen G, Wittfoth M, Frieling H, et al. The Neurobiology and Psychology of Pedophilia: Recent Advances and Challenges. Front Hum Neurosci 2015; 9: 344. 2015/07/15. DOI: 10.3389/fnhum.2015.00344.

7. Mohnke S, Muller S, Amelung T, et al. Brain alterations in paedophilia: a critical review. Prog Neurobiol 2014; 122: 1–23. 2014/08/15. DOI: 10.1016/j.pneurobio.2014.07.005.

8. Poeppl TB, Nitschke J, Santtila P, et al. Association between brain structure and phenotypic characteristics in pedophilia. J Psychiatr Res 2013; 47: 678-685. 2013/02/13. DOI: 10.1016/j.jpsychires.2013.01.003.

9. Cantor JM, Kabani N, Christensen BK, et al. Cerebral white matter deficiencies in pedophilic men. J Psychiatr Res 2008; 42: 167-183. 2007/11/28. DOI: 10.1016/j.jpsychires.2007.10.013.

10. Schiffer B, Paul T, Gizewski E, et al. Functional brain correlates of heterosexual paedophilia. Neuroimage 2008; 41: 80-91. 2008/03/25. DOI: 10.1016/j.neuroimage.2008.02.008.

11. Keshavan MS and Kaneko Y. Secondary psychoses: an update. World Psychiatry 2013; 12: 4–15. 2013/03/09. DOI: 10.1002/wps.20001.

12. McAllister TW. Neurobehavioral sequelae of traumatic brain injury: evaluation and management. World Psychiatry 2008; 7: 3–10. 2008/05/07. DOI: 10.1002/j.2051-5545.2008.tb00139.x.

13. Camperio Ciani AS, Scarpazza C, Covelli V, et al. Profiling acquired pedophilic behavior: Retrospective analysis of 66 Italian forensic cases of pedophilia. Int J Law Psychiatry 2019; 67: 101508. 2019/12/02. DOI: 10.1016/j.ijlp.2019.101508.

14. Gilbert F and Focquaert F. Rethinking responsibility in offenders with acquired paedophilia: punishment or treatment? Int J Law Psychiatry 2015; 38: 51-60. 2015/03/03. DOI: 10.1016/j.ijlp.2015.01.007.

15. Gomes Lopez PM, de Castro Prado CS and de Oliveira-Souza R. The Neurology of Acquired Pedophilia. Neurocase 2020; In press.

16. Scarpazza C, Pellegrini S, Pietrini P, et al. The role of Neuroscience in the Evaluation of Mental Insanity: on the controversies in Italy. Neuroethics 2018; 11: 83–95.

17. Burns JM and Swerdlow RH. Right orbitofrontal tumor with pedophilia symptom and constructional apraxia sign. Arch Neurol 2003; 60: 437-440. 2003/03/14.

18. Sartori G, Scarpazza C, Codognotto S, et al. An unusual case of acquired pedophilic behavior following compression of orbitofrontal cortex and hypothalamus by a Clivus Chordoma. J Neurol 2016; 263: 1454-1455. 2016/05/11. DOI: 10.1007/s00415-016-8143-y.

19. von Krafft-Ebing B. Trattato di Psicopatologia Forense. Fratelli Bocca, Editori (Torino) 1897: 215.

20. Mendez M and Shapira JS. Pedophilic behavior from brain disease. J Sex Med 2011; 8: 1092-1100. 2011/01/18. DOI: 10.1111/j.1743-6109.2010.02172.x.

21. Miller BL, Cummings JL, McIntyre H, et al. Hypersexuality or altered sexual preference following brain injury. J Neurol Neurosurg Psychiatry 1986; 49: 867-873. 1986/08/01. DOI: 10.1136/jnnp.49.8.867.

22. Eickhoff SB, Bzdok D, Laird AR, et al. Activation likelihood estimation meta-analysis revisited. Neuroimage 2012; 59: 2349-2361. 2011/10/04. DOI: 10.1016/j.neuroimage.2011.09.017.

23. Darby RR, Horn A, Cushman F, et al. Lesion network localization of criminal behavior. Proc Natl Acad Sci U S A 2018; 115: 601-606. 2017/12/20. DOI: 10.1073/pnas.1706587115.

24. Moher D, Liberati A, Tetzlaff J, et al. Preferred reporting items for systematic reviews and meta-analyses: the PRISMA statement. BMJ 2009; 339: b2535. 2009/07/23. DOI: 10.1136/bmj.b2535.

25. Müller VI, Cieslik EC, Laird AR, et al. Ten simple rules for neuroimaging meta-analysis. Neuroscience Biobehavioral Reviews 2018; 84 151–161.

26. Turkeltaub PE, Eickhoff SB, Laird AR, et al. Minimizing within-experiment and within-group effects in Activation Likelihood Estimation meta-analyses. Human Brain Mapping 2012; 33: 1–13.

27. Eickhoff SB, Stephan KE, Mohlberg H, et al. A new SPM toolbox for combining probabilistic cytoarchitectonic maps and functional imaging data. Neuroimage 2005; 25: 1325–1335. 2005/04/27. DOI: 10.1016/j.neuroimage.2004.12.034.

28. Rorden C, Fridriksson J and Karnath HO. An evaluation of traditional and novel tools for lesion behavior mapping. Neuroimage 2009; 44: 1355-1362. 2008/10/28. DOI: 10.1016/j.neuroimage.2008.09.031.

29. Genon S, Reid A, Langner R, et al. How to Characterize the Function of a Brain Region. Trends Cogn Sci 2018; 22: 350-364. 2018/03/05. DOI: 10.1016/j.tics.2018.01.010.

30. Eickhoff SB, Nichols TE, Laird AR, et al. Behavior, sensitivity, and power of activation likelihood estimation characterized by massive empirical simulation. Neuroimage 2016; 137: 70-85. 2016/05/18. DOI: 10.1016/j.neuroimage.2016.04.072.

31. Fagan PJ, Wise TN, Schmidt CW, Jr., et al. Pedophilia. JAMA 2002; 288: 2458-2465. 2002/11/19. DOI: 10.1001/jama.288.19.2458.

32. Devinsky J, Sacks O and Devinsky O. Kluver-Bucy syndrome, hypersexuality, and the law. Neurocase 2010; 16: 140–145. 2009/11/21. DOI: 10.1080/13554790903329182.

33. Scarpazza C, Pennati A and Sartori G. Mental Insanity Assessment of Pedophilia: The Importance of the Trans-Disciplinary Approach. Reflections on Two Cases. Front Neurosci 2018; 12: 335. 2018/06/06. DOI: 10.3389/fnins.2018.00335.

34. Regestein QR and Reich P. Pedophilia occurring after onset of cognitive impairment. J Nerv Ment Dis 1978; 166: 794-798. 1978/11/01.

35. Frohman EM, Frohman TC and Moreault AM. Acquired sexual paraphilia in patients with multiple sclerosis. Arch Neurol 2002; 59: 1006-1010. 2002/06/12.

36. Fumagalli M, Pravettoni G and Priori A. Pedophilia 30 years after a traumatic brain injury. Neurol Sci 2015; 36: 481–482. 2014/08/08. DOI: 10.1007/s10072-014-1915-1.

37. Raymond NC, Coleman E, Ohlerking F, et al. Psychiatric comorbidity in pedophilic sex offenders. Am J Psychiatry 1999; 156: 786-788. 1999/05/18. DOI: 10.1176/ajp.156.5.786.

38. Anckarsäter H, Radovic S, Svennerlind C, et al. Mental disorder is a cause of crime: the cornerstone of forensic psychiatry. Int J Law Psychiatry 2009; 32: 342–347.

39. Crossley NA, Scott J, Ellison-Wright I, et al. Neuroimaging distinction between neurological and psychiatric disorders. Br J Psychiatry 2015; 207: 429-434. 2015/06/06. DOI: 10.1192/bjp.bp.114.154393.

40. McGorry P, Keshavan M, Goldstone S, et al. Biomarkers and clinical staging in psychiatry. World Psychiatry 2014; 13: 211-223. 2014/10/03. DOI: 10.1002/wps.20144.

